# The role of rare genetic variants enrichment in epilepsies of presumed genetic etiology

**DOI:** 10.1101/2023.01.17.23284702

**Authors:** Linnaeus Bundalian, Yin-Yuan Su, Siwei Chen, Akhil Velluva, Anna Sophia Kirstein, Antje Garten, Saskia Biskup, Florian Battke, Dennis Lal, Henrike O. Heyne, Konrad Platzer, Chen-Ching Lin, Johannes R. Lemke, Diana Le Duc, Epi25 Collaborative

## Abstract

Previous studies suggested that severe epilepsies e.g., developmental and epileptic encephalopathies (DEE) are mainly caused by ultra-rare *de novo* genetic variants. For milder phenotypes, rare genetic variants could contribute to the phenotype. To determine the importance of rare variants for different epilepsy types, we analyzed a whole-exome sequencing cohort of 9,170 epilepsy-affected individuals and 8,436 controls. Here, we separately analyzed three different groups of epilepsies : severe DEEs, genetic generalized epilepsy (GGE), and non-acquired focal epilepsy (NAFE). We required qualifying rare variants (QRVs) to occur in controls at a minor allele frequency ≤ 1:1,000, to be predicted as deleterious (CADD≥20), and to have an odds ratio in epilepsy cases ≥2. We identified genes enriched with QRVs in DEE (n=21), NAFE (n=72), and GGE (n=32) - the number of enriched genes are found greatest in NAFE and least in DEE. This suggests that rare variants may play a more important role for causality of NAFE than in DEE. Moreover, we found that QRV-carrying genes e.g., *HSGP2, FLNA* or *TNC* are involved in structuring the brain extracellular matrix. The present study confirms an involvement of rare variants for NAFE, while in DEE and GGE, the contribution of such variants appears more limited.

## 2. Introduction

Epilepsy is one of the most common neurological diseases worldwide, affecting almost 1% of the population in the United States ^1^. Early pedigree studies showed a high genetic component and a heritability of up to 70% ^2,3^. With the help of next generation sequencing there was a significant advance in gene discovery. Currently, hundreds of genes are established as monogenic causes for epilepsy ^4^, while recent studies have associated a few to polygenic causes ^5^. Yet, the biggest leap in diagnostic yield happened mainly for the most severe type of epilepsies, developmental and epileptic encephalopathy (DEE) ^6^. For this type of epilepsy, the heritability or susceptibility are very low, since such diseases are often caused by deleterious *de novo* variants as the severely affected individuals usually do not reproduce.

The role of common ^7–9^ and ultra-rare *de novo* genetic variants ^10,11^ for epilepsy has been extensively researched. Epidemiological studies accounting for the similar prevalence across populations and the increased risk of individuals in more densely affected families, suggested that polygenic predisposition should have a predominant role over the monogenic etiology ^12^. This has been addressed by genome wide association studies and polygenic risk scores ^7–9^ which identified mostly non-coding variants with individually small effects – median odds ratio (OR) generally lower than 1.3, but with a high aggregate effect explaining in part the missing heritability. Conversely, the ultra-rare *de novo* genetic variants have much larger effects on individual risk, but they make only a small contribution to the overall heritability in the population owing to their rarity ^13,14^.

Our understanding of the underlying genetic architecture leading to increased susceptibility to epilepsy due to a middle tier of variants that are rare (neither ultra-rare *de novo*, nor common, i.e. allele frequency of > 1%) is still very limited. Based on evolutionary theory, forces of negative natural selection will keep large-effect risk variants at much lower frequencies in the population, especially for a disorder like epilepsy which results in reduced fitness, i.e., reproduction. Analysis of rare variants’ contribution to the disease could be a useful tool for a better understanding of the heritability and disease pathomechanism ^15^ as it was shown in some other conditions e.g., autism.^16^

In this study, focused only on rare variants (minor allele frequency ≤ 1:1,000) predicted to be deleterious and with an excess in cases (OR ≥ 2), which best reflects the effect size ^15^. Finally, to understand the underlying pathomechanism we performed a combined analysis of the identified genes and their interacting partners aimed at identifying molecular pathways, which are potentially disrupted.

## 3. Materials and Methods

### 3.1. Cohort and data description

Genetic and phenotype information were obtained from the Epi25 collaborative ^11^ (http://epi-25.org/). Phenotyping procedures, case definitions, and ancestry of the participating individuals are reported in a previous Epi25 collaborative study ^11^. To account for differences in ancestry and exome capture technologies among individuals the data has undergone previously described thorough quality check procedures and only individuals of European descent were analyzed ^11^. Briefly, variant calling was performed with GATK ^17^ and only variants with a genotype quality > 20 were kept. Variants called heterozygous were required to have an allele frequency of 0.2–0.8. To control for kit enrichment artefacts, only variants where 80% of both Agilent and Illumina-sequenced samples show at least 10-× coverage were retained. Ancestry stratification had previously been ruled out using principal-component analyses to identify ancestral backgrounds and only individuals of European ancestry classified by Random Forest with 1,000 Genomes data were further analyzed. Annotation of variants was performed with Ensembl’s Variant Effect Predictor ^18^ for human genome assembly GRCh37.

To understand differences in genetic susceptibility across different types of epilepsy, we analyzed 1, 021 individuals with developmental and epileptic encephalopathy (DEE), 3, 108 individuals with genetic generalized epilepsy (GGE), and 3, 597 individuals with non-acquired focal epilepsy (NAFE). Each cohort was compared to 8, 436 matched-ancestry, unrelated controls.^11^

### 3.2 Qualifying rare variants and variant set enrichment analysis

We defined a variant as a qualifying rare variant (QRV) if it met following criteria:

- The variant is present in controls i.e. AC_CTRL ≥ 1;
- The minor allele frequency ≤ 1:1,000;
- The variant is predicted to be deleterious – CADD score ≥ 20 ^19^;
- OR ≥ 2, *p*-value ≤ 0.05 in epilepsy cases.

To account for gene length and different mutation rate across genes, we counted the total number of observed variants per gene and the number of variants with AF ≤ 0.001. Using these two parameters, we modeled a simple linear regression to characterize the relationship of the number of rare variants and total variants per gene. Based on the linear regression, we estimated the expected number of rare variants with OR ≥ 2 for each gene and compared to the observed. Only genes with an excess of rare variants were considered in the QRV filtering (Supplemental Fig. 1).

The susceptibility and risk burden for each gene were estimated by testing for enrichment of QRVs. To this end, we assigned an empirical enrichment score (ES) to each gene. The generation of the ES was inspired by the Significance and Functional Enrichment (SAFE) framework^20^ and Gene Set Enrichment Analysis (GSEA) hence termed as Variant Set Enrichment Analysis (VSEA).

VSEA allows us to score the genes based on the number of QRVs they have across the general population and epilepsy patients. We defined following lists of variants:

- ranked variant list (L) – list of QRVs per epilepsy type ordered (descending) by their corresponding OR;
- background set (S) – list of variants grouped into synonymous or nonsynonymous variants per epilepsy type.

The enrichment analysis is designed to check if variants in S are randomly distributed throughout L, or skewed to a side where OR is higher or lower. With this method, we can also identify the variants in each gene which contribute most to the enrichment – referred to as leading edge.

The enrichment score (ES) assigned to the genes is calculated using the maximum deviation observed among cumulative ranked sum ^20^ of both ***hits*** and ***miss*** across the variants (Supplemental Fig. 2) normalized by the number of observed ***hits and miss***. The vector of either ***hits*** or ***miss*** can be represented by vector *y* and ES as the maximum difference between the two vectors, defined as follow:

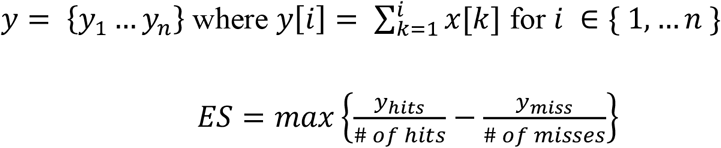

A high ES of a gene indicates an enrichment of variants having higher OR in comparison to the variants found in the lower rank. After calculating the ES, to determine the significance we performed n = 1,000 permutations where a set of variants having the length *n(L)* were randomly selected from set *S*. This was used to calculate an empirical distribution of ES. The number of times the empirical ES exceeds the observed ES was counted and divided by the number of permutations (n) to calculate the *p*-values (pval).

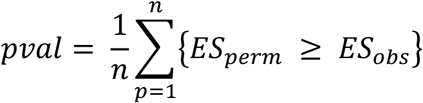

For multiple testing correction, we determined FDR using the ratio of the Normalized Enrichment Score (NES) – observed and permuted ES. NES is the ES divided by the expected ES i.e., average ES from the permuted values.

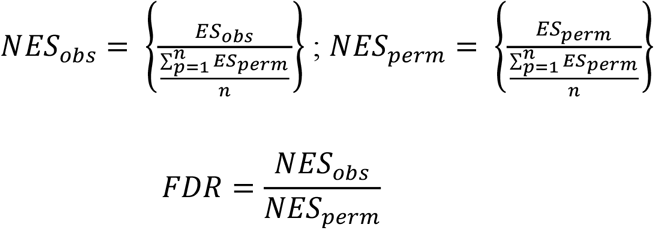

### 3.3 Functional module analysis and protein-protein interactions

To test in which functional pathway the genes identified that bear an excess of QRVs play a role, we performed enrichment analysis across multiple gene sets including but not limited to Gene Ontology (GO), Allen Brain Atlas, Reactome, KEGG pathways.

#### 3.3.1 Allen Brain Atlas Enrichment

To identify gene sets that are specific to certain brain areas and/or developmental stages we used the Allen Human Brain Atlas. This resource delivers information about gene expression levels in various parts of the human brain during the course of brain development ^21,22^. We used the R package *ABAEnrichment* to test whether genes with excess QRVs show significant enrichment in specific brain regions or brain developmental stages ^23^.The package integrates human brain expression datasets provided by the Allen Brain Atlas in both the prenatal and adult stage. The expression data is analyzed over 47 brain regions and 20 age time points. The gene expression is evaluated during development from prenatal stage to adult. If the change is high in a specific region, the gene is annotated to that region and the score mirrors the deviation from prenatal to adult stage ^23^.

#### 3.3.2 Overrepresentation Analysis (ORA)

The gene lists were also subjected to functional class analysis using ORA with MsigDb Reactome database for gene set collections C2 (Reactome) and C5 (Gene Ontology) ^24,25^ and GOFuncR ^26^. The method uses a hypergeometric test ^27^ to assess the probability of observing at least *k* genes from the list across the pathway database ^24,28^.

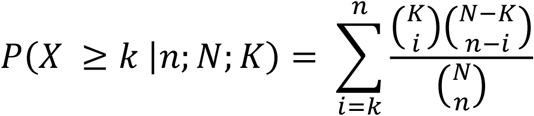

For the customized gene set ORA, we used the datasets from MSigDB – C5 for sets associated with ion channels, neurotransmitter, glutamatergic and GABAergic signaling, nervous system development, and synaptic functions ^24,25^. For GoFuncR overrepresentation, we used C5 (Gene Ontology) of MSigDB for restricting the background genes to those which are found expressed in the brain ^29^.

#### 3.3.3 Analysis of distance in the human protein interacting network (PIN)

To elucidate the putative roles of the genes significantly enriched with QRVs in epilepsy, we investigated the distance between them and the known epilepsy-related genes in the human PIN. The source of the human PIN data is InBio Map ^30^ and epilepsy-related genes were acquired from the consolidated list of epi-25.org. The distance is defined as the shortest path length between gene *u* and *v* in the human PIN. All paired shortest path lengths between genes are calculated by the Dijkstra algorithm. We identified which genes with QRVs are significantly closer located to the known epilepsy-related genes within the PIN, compared to the distance of all other genes.

## 4. Results

### 4.1 Enrichment of QRVs in three types of epilepsies

We tested the burden of QRVs per gene in each epilepsy group: DEE, NAFE, and GGE. The only significant gene which was present across all types of epilepsies was *HSPG2* (*p*-value = 0.0001, from 10,000 random samplings). *HSPG2* (Heparan Sulfate Proteoglycan 2) encodes the perlecan protein that belongs to the glycosaminoglycans family, which are major components of the brain extracellular matrix (ECM). Although the gene was common to all epilepsy groups, there were different variants in this gene, which contributed to the increased enrichment score of the gene for each group (Supplementary Table 1).

Further, consistent with the presumed *de novo* pathogenic variants occurrence and a highly penetrant phenotype, DEE showed the lowest number of genes enriched with QRVs (Fig. 1A); this supports DEE’s mainly monogenic origin. In other words, for DEE, the most severe of the epilepsy phenotypes, it is less likely that variants present in controls contribute as causal risk factors. The largest number of genes with a significantly high enrichment score was retrieved for NAFE (Fig. 1A, Supplemental Table 1). This result contrasts the previous findings on ultra-rare *de novo* genetic variants, which show mainly no significant burden in the NAFE individuals ^11^ and may suggest that rare variants also present in controls could contribute more to the NAFE pathophysiology compared to highly damaging *de novo* variation.

**Fig. 1.**
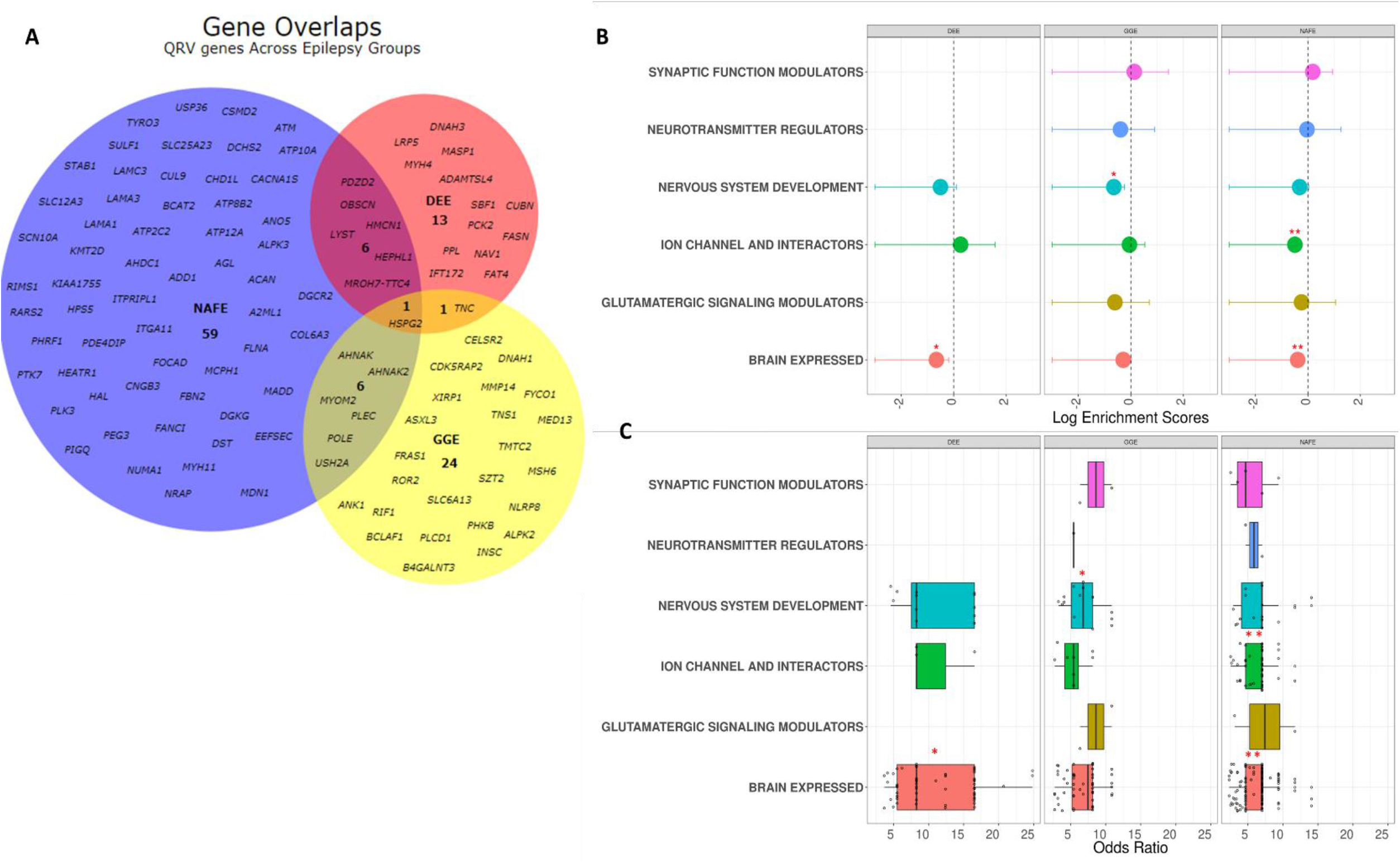
**A. QRV enriched genes across different epilepsy groups.** For each epilepsy group, a set of genes were identified to be enriched with QRVs (FDR ≤ 0.05, Z score ≥ 2). The number of genes found for NAFE are seemingly greater than the two other groups implying that rare variants in this particular group could have a larger contribution to the etiology compared to DEE and GGE. **B. Overrepresentation of QRV enriched genes**. The graph shows the log enrichment scores and intervals with confidence interval = 0.95 for the overrepresentation of the QRV enriched genes. The genes enriched with QRVs were found to be overrepresented across highly expressed genes in the brain (DEE, NAFE), nervous system development (GGE), and ion channel and interactors (NAFE) – suggesting a role in the brain and brain-related processes. C. Odds ratio distribution of QRV enriched genes across different gene sets. The graph shows the odds ratio distribution of the variants per genes across different gene sets. (* FDR ≤ 0.05, ** FDR ≤ 0.01)

To gain insight into the functionality of the genes, we classified them into 6 categories, which are considered to play a role in the epileptic pathomechanism: modulators of synaptic functions, neurotransmitter regulators, nervous system development, ion channels and their interacting partners, modulators of glutamatergic signaling, and genes with high expression in brain (Fig. 1 B, C, Table 1). Our results showed for DEE (FDR = 0.03)and NAFE (FDR = 0.004) a significant enrichment especially for genes highly expressed in brain. GGE showed an enrichment for genes annotated to nervous system development (FDR = 0.026). In NAFE we identified genes enriched for ion channels and their interactors (FDR = 0.004) (Fig. 1B). We showed that variants in these genes generally had median ORs > 5 (Fig. 1C), suggesting they may have a large effect size and could contribute to the underlying pathomechanism.

**Table 1.**
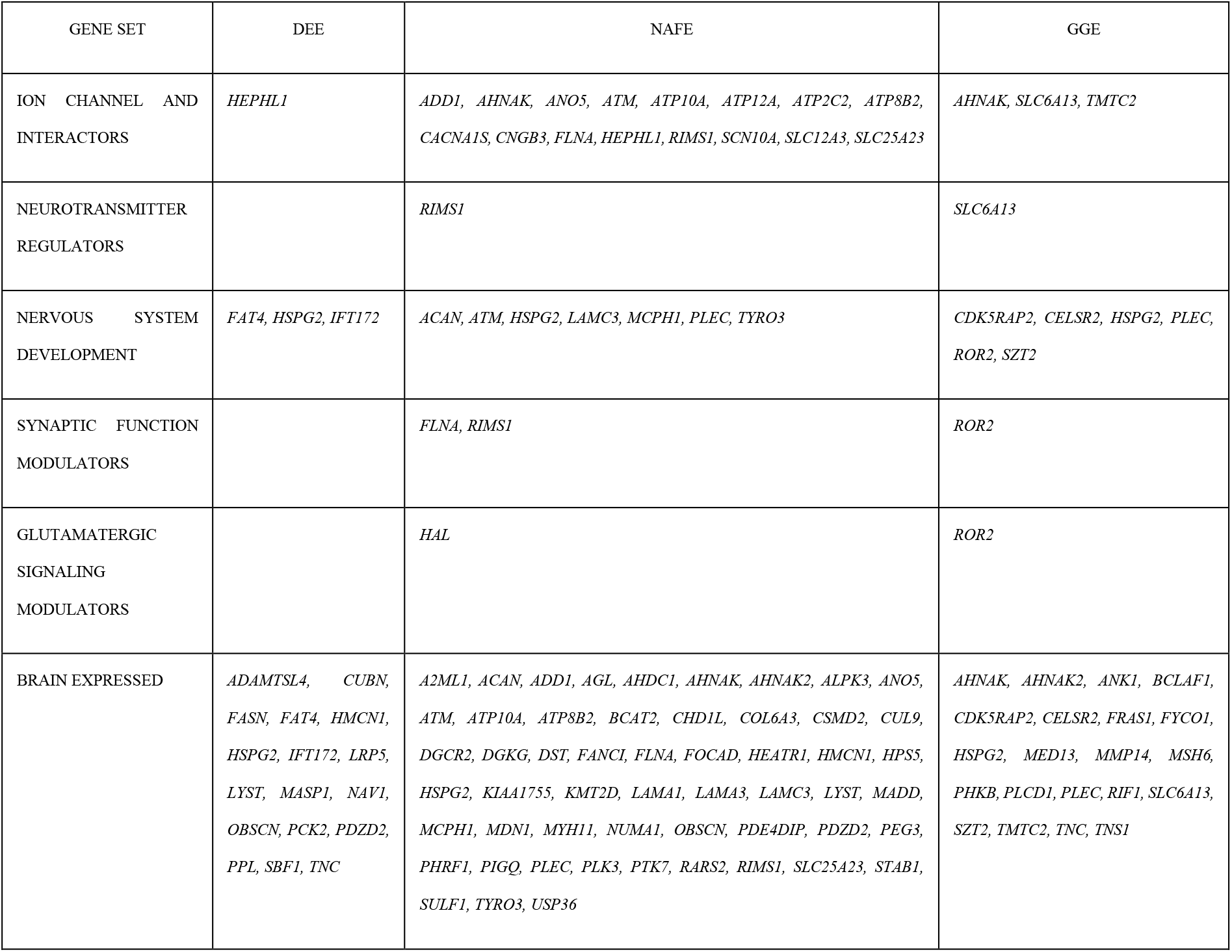
Overrepresented genes across gene sets annotated to brain processes and molecular functions. The table contains the QRVs-enriched genes, which are found to be enriched in gene sets derived from molecular functions in GO database and brain gene expression level from Protein Atlas and GTEx. For each gene set, we selected processes associated with ion channels, GABAergic and glutamatergic related pathways, synaptic functions, neurotransmission, nervous system development and genes with nTPM ≥ 1 in brain. ^29^

### 4.2 QRVs collapsing captures genes encoding for ion channels or their interactors in NAFE

Ion channels play a major role in genetic epilepsies ^31,32^. Although, since the beginning of the epilepsy-related gene discoveries, many other gene classes and biological pathways have been revealed to play a role, it is still a significant proportion (∼ 25 %) of the epilepsy genes known to date that encode for ion channels ^31^. Since we identified the ion channel-related molecular function pathway to be enriched in NAFE (Fig. 1B), we performed ORA based on KEGG and Reactome ^33^. For KEGG, the ion channel pathway is annotated only with respect to drug development, and thus, we could not identify an over-representation based on the QRVs-enriched genes in NAFE. Using the Reactome annotation we identified the ion channel transport category to be significantly enriched. NAFE-related genes annotated to the Reactome category are: *ATP2C2, ATP12A, ATP8B2, ATP10A*, and *ANO5*. While, the first four genes encode for ATPases involved in ions transport, like Ca^2+^ or H^+^/K^+^, *ANO5* encodes for an anoctamin, which belongs to a protein family of Ca^2+^ activated chlorine channels and phospholipid scramblases ^34^. A founder mutation in *ANO5* has been implicated in muscular dystrophy ^34^. Despite the high expression in brain and the controversial muscle phenotype in the mouse knockout models ^35,36^, its function in the brain remains unknown.

Using the gene ontology annotation, we identified a few ion channel genes and multiple genes interacting with ion channels, which showed QRVs enrichment in NAFE (Table 1), e.g. *CACNA1S* and *SCN10A*. Another identified gene is *ADD1* coding for adducin. Although adducin is primarily responsible for the assembly of spectrin-actin that provides functional support to the cytoskeleton, the gene ontology also annotates the gene to ion transport and synaptic functions. Variants in *ADD1* have also been recently identified in intellectual disability, corpus callosum dysgenesis, and ventriculomegaly in humans ^37^. Similarly, ATM, another gene with QRVs for NAFE, has been recently shown to be involved in hippocampal and cortical development, as well as synaptic functions ^38^.

Based on our analysis, we did not retrieve genes encoding for ion channels that have already been associated with monogenic epilepsy. Those genes generally bear ultra-rare variants that do not occur in controls, which does not comply with our definition of QRVs. Some of the genes we identified to be significantly enriched in NAFE have already been associated with mendelian disorders e.g., *HSPG2* and *FLNA*, yet their phenotype does not appear to be severe or highly penetrant which meets our hypothesis that variants in these genes could confer an increased risk for epilepsy.

### 4.3 Involvement in brain development

We asked whether the genes with QRVs are involved in brain development and in which brain regions they are most relevant. Patterns of gene expression can be very informative in respect to the importance of a gene during development. Using the ABAEnrichment package ^23^, we tested whether the genes identified in the different types of epilepsy play a role during development in any brain regions. From the results, we identified genes from DEE to show the highest involvement during development with significant enrichment over 13 brain regions (Fig. 2). For GGE only the cerebellar cortex showed a signal (family wise error rate, FWER = 0.045), while for NAFE we identified a signal in this region and two additional ones in the striatum and inferolateral temporal cortex. When we inquired the developmental scores assigned based on expression changes between prenatal and adult stage, we identified genes encoding for ECM proteins (*LAMA1, FBN2, COL6A3*) to contribute to the enrichment in the different brain regions for NAFE (Fig. 3). Similarly, *TNC*, which encodes for the ECM protein tenascin C showed a high developmental score contributing to the brain regions enrichment in both DEE and GGE. For DEE, *FAT4*, a gene encoding for a protocadherin, a calcium-dependent cell adhesion protein, showed the highest developmental score (Fig. 3). *FAT4*, which was previously related to epilepsy ^39^, plays a role in the maintenance of planar cell polarity as well as in neuroprogenitor proliferation ^40^. For GGE, *ROR2* is the leading gene in respect to the developmental score. *ROR2* encodes a tyrosine-protein kinase transmembrane receptor also known as the neurotrophic tyrosine kinase, receptor-related 2, which also appears to play a role in the maintenance of neuroprogenitor cells in the developing neocortex ^41^. Based on the observed functions for the genes with the highest developmental scores, we were further prompted to perform pathway analyses and understand in which molecular processes genes with QRVs are involved.

**Fig. 2.**
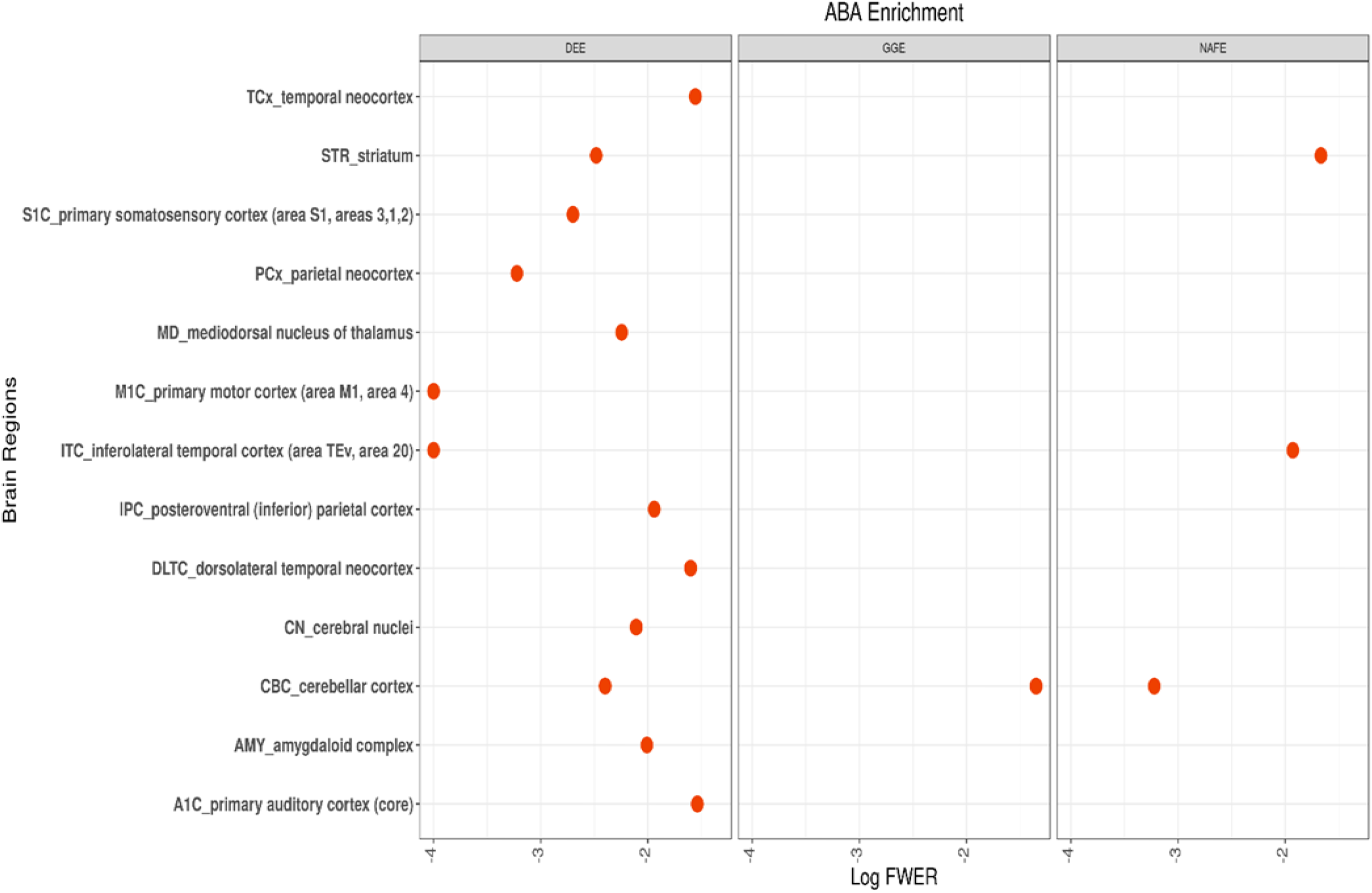
A. ABA Enrichment of QRV enriched genes across all epilepsy types. The graph shows brain regions with enriched QRVs genes with respect to their developmental score and the log transformed FWER (Family wise error rate) associated with the significance of the enrichment.

**Fig. 3.**
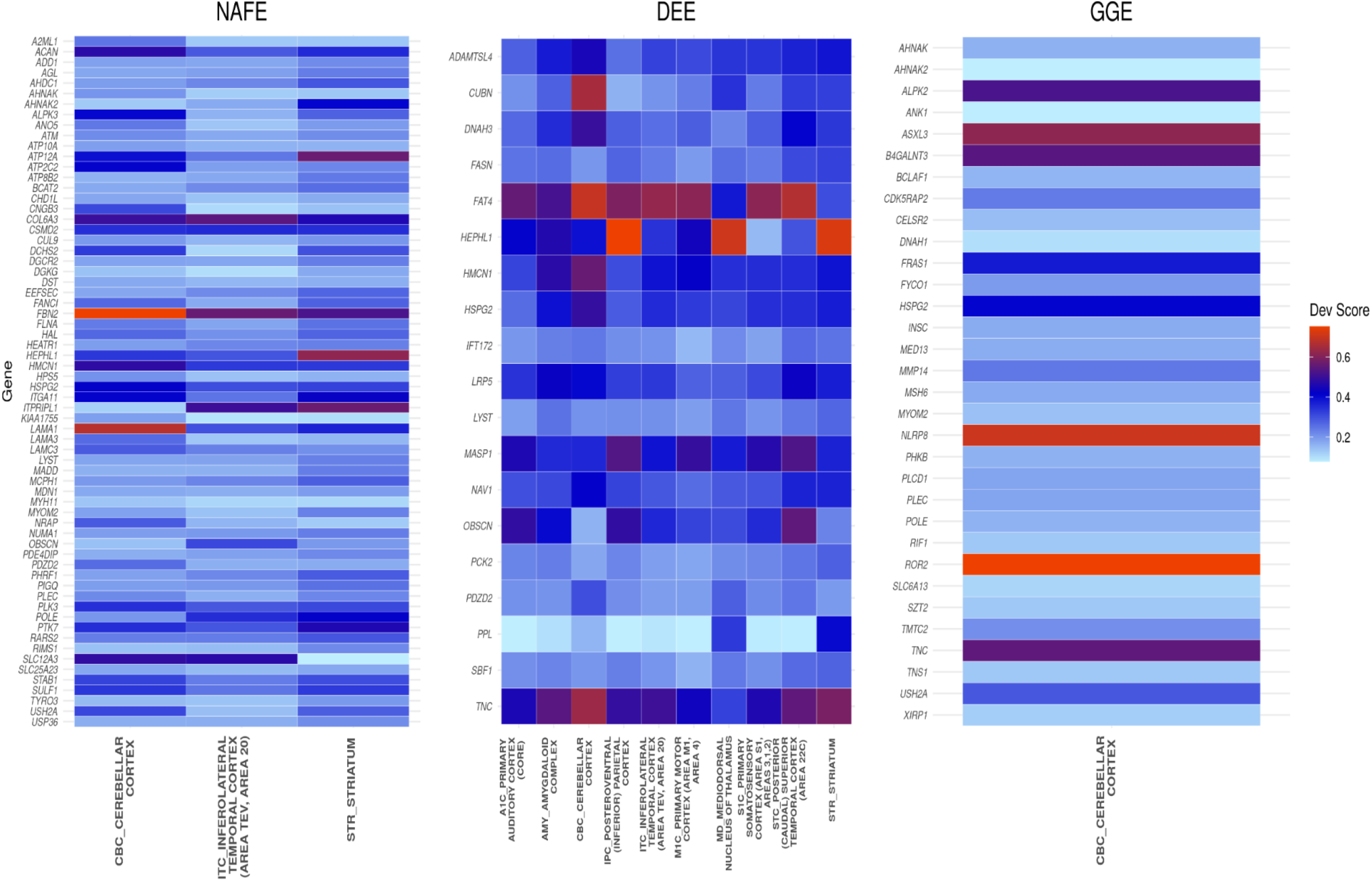
ABA Developmental score of QRV enriched genes. Heatmap with developmental scores of QRV genes significantly associated with the different brain regions during development. The listed regions were identified to be overrepresented with QRV enriched genes.

### 4.4 Pathway and network analyses

We performed pathway enrichment analyses to determine whether the identified genes cluster within specific functions. For DEE we could not identify any significant categories after multiple testing correction. NAFE showed enrichment of many GO categories, that clustered mainly within the extracellular matrix (ECM) or cell adhesion (Fig. 4A). For GGE, there were only 3 significant categories, all related to cellular junctions or adhesion (Supplemental Table 2).

**Fig. 4.**
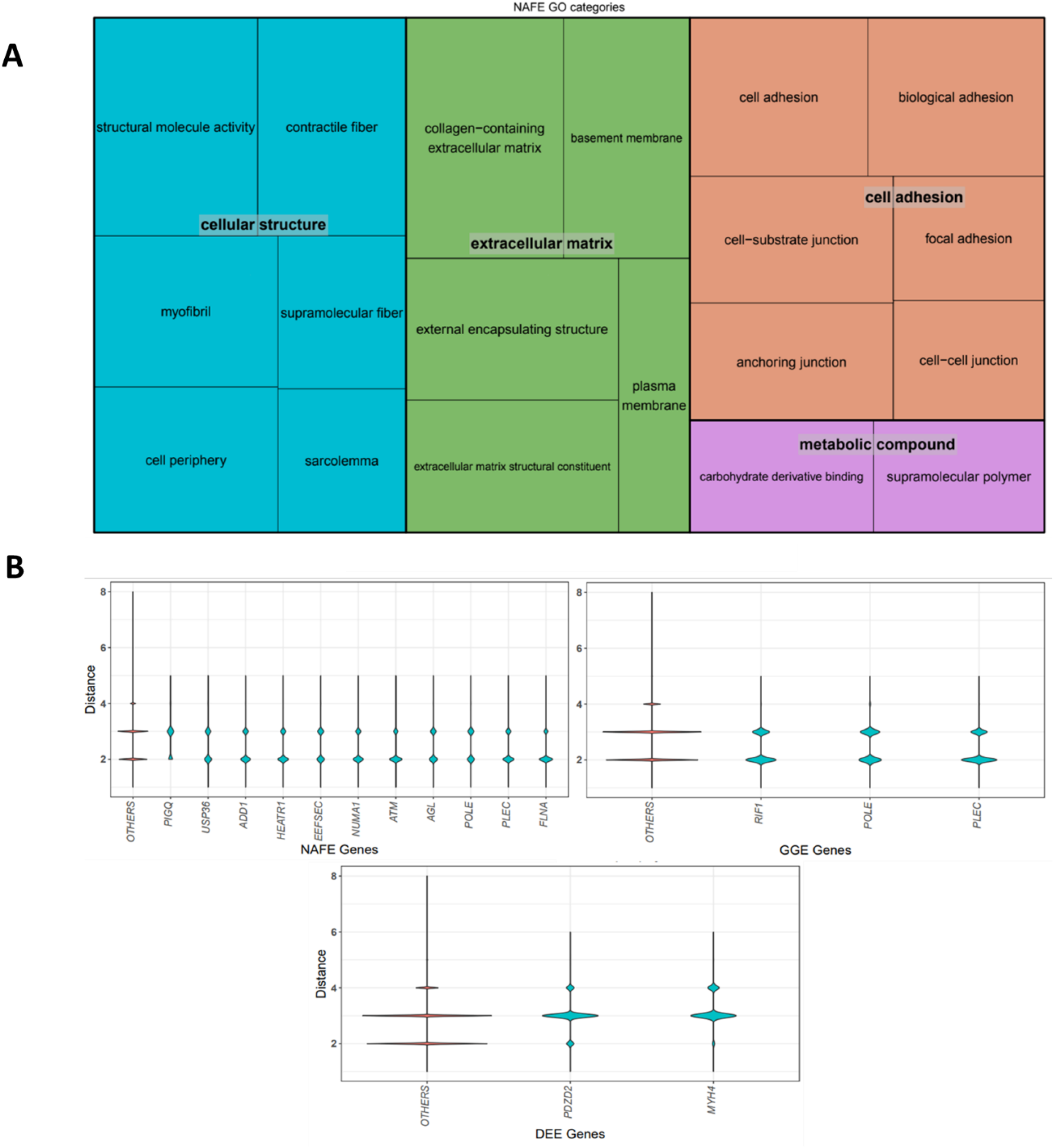
**A. Overrepresentation of QRV genes from NAFE.** The GO terms overrepresented by the genes found to be enriched in NAFE cluster into a more general category to represent their functional class. The overrepresentation analysis was done using GoFuncR with a gene expressed in brain as background. **B. Genes and their distance to known epilepsy genes**. The distance (y-axis) of the QRV genes (x-axis) to known epilepsy genes was determined by counting the nodes within the protein-protein interaction network. We show genes with significantly shorter distance to known epilepsy genes compared to the distance of randomly selected genes. A larger distribution assigned to the lower distance in the violin plot implies a higher number of genes with short distance to known epilepsy genes in the PIN. The short distance to the epilepsy genes could suggest a contribution to epilepsy pathomechanism.

To further test how the identified genes with QRVs are connected to already known epilepsy genes, we determined the distance between the identified gene and the epilepsy genes from Epi25 (http://epi-25.org/). To this end, we assessed within the PPI network how many nodes represented by protein interacting partners lie between the identified gene and any known epilepsy gene. While for NAFE, the mean distance between QRV genes and epilepsy genes was at 2.57, significantly smaller than the mean distance of between the non-QRV genes and epilepsy genes (2.66, *p*-value < 0.001), the effect size is too small to conclude that overall, the identified genes with QRVs are closer to known epilepsy genes. We thus, further determined the genes with a shorter distance to known epilepsy genes within the PIN (Fig. 4B). For GGE and DEE the overall mean distance did not reach significance and only 3 and 2 genes, respectively, showed a significantly closer distance to epilepsy genes.

## 5. Discussion

Unlike common and *de novo* ultra-rare variants, rare variants in epilepsy have not been researched extensively. Based on the ‘Common Disease, Rare Variant’ hypothesis multiple rare sequence variants, with relatively high penetrance, confer an increased genetic susceptibility to a common disease ^42^. While the identification of such variants is paramount for understanding their role, their detection is statistically more challenging because they are present at low frequencies in the general population. To understand how rare variants contribute to the etiology and pathomechanism of epilepsy, we applied a method to estimate the risk burden of a gene based on the enrichment of rare variants posing a relatively high risk in the Epi25 cohort i.e., OR ≥ 2. Using the proportion of QRVs in cases and controls, we calculated a score for each gene. A high score of a gene implies a higher disease probability due to the presence of the rare variants with higher ORs in cases. By this approach we identified sets of QRV enriched genes – DEE (n = 21), NAFE (n = 72), and GGE (n = 32) (Fig. 1). In a previous study that analyzed the same patient cohort in respect to the enrichment of deleterious ultra-rare variants, DEE and GGE individuals had significantly more such variants compared to those diagnosed with NAFE ^11^. While for our study on the enrichment of rare variants, we identified the lowest number of genes for DEE and the highest for NAFE. Our results and the results of the previous study ^11^ could suggest that, while for DEE the pathomechanism relies on highly deleterious and penetrant variants, NAFE may result from an enrichment of more frequent and less penetrant rare variants, identified also in controls, albeit at lower frequency than in cases.

To further analyze the gene set and its relevance to specific molecular pathways, we performed a GO enrichment analysis. This revealed a significant overrepresentation of the NAFE genes across ECM and structural related pathways (Fig. 4A). The ECM is known to play a role in epileptogenesis since it is involved in the establishment of neural plasticity, i.e. cell-cell connections and signaling ^43^. Moreover, changes in ECM have been directly implicated in the pathophysiology of temporal lobe epilepsy ^44,45^, the most common form of focal epilepsies/NAFE. Interestingly, *HSPG2*, the only gene found to be enriched across all types of epilepsy in the study, encodes for perlecan, an important member of brain ECM. Little is known about *HSPG2* and its association to epilepsy, but some studies have revealed its role in acetylcholinesterase clustering at the synapse, which has the capability to interfere in synaptic transmission ^46^. Perlecan is, however, ubiquitously expressed and pathogenic variants have been implicated in the Schwartz–Jampel syndrome type I, a rare autosomal recessive disease with cardinal symptoms consisting of skeletal dysplasia and neuromuscular hyperactivity ^47^. Some of the affected individuals also show impaired neurologic development, consistent with perlecan’s neuroprotective effect and its involvement in neurogenesis and normalization of neocortical excitability after insult events ^48^. In further support of our finding, previous studies have also considered *HSPG2* to be an epilepsy-associated gene, although the underlying mechanism is still not clear ^39^.

While among the identified genes, there was an enrichment of genes with high brain expression, both in the DEE and NAFE groups, for NAFE we additionally identified variants in ion channels and their interactors to play a role (Fig. 1 B, C). *CACNA1S* is one of the genes with an excess of QRVs in NAFE individuals. The gene is lowly expressed in the brain and highly expressed in the muscles being implicated in the hypokalemic periodic paralysis. However, we observed outliers in respect to brain expression (Supplemental Fig. 3) ^49^, suggesting that Cav1.1, the L-type voltage gated calcium channel encoded by *CACNA1S*, could play a role in the calcium influx in response to large depolarizing shifts in membrane potential for some individuals. In support of the variability of the *CACNA1S*’ brain involvement, rare variants in this gene have been associated with schizophrenia ^50^. Similarly, *SCN10A* is lowly expressed in the brain, but shows an enrichment signal in our dataset. Biallelic variants in this gene have been potentially linked to epilepsy-related phenotypes ^51^.

In an additional analysis, to understand whether the genes we identified are closer in the PIN to already known epilepsy genes, we calculated the PPI distance (Fig. 4B). Most of the genes with significantly shorter PPI paths connect them to epilepsy genes for NAFE. A gene with significantly shorter distance to known epilepsy genes is *PDZD2*, enriched for both NAFE (FDR = 0.005) and DEE (FDR = 0.006). This gene can be found expressed mainly in the basal ganglia and cerebral cortex with high specificity among oligodendrocytes precursor cells, and excitatory neurons ^28,51^. *PDZD2* also contributes to the functional expression of Nav1.8 ion channel, which is encoded by *SCN10A*, a QRV enriched gene in NAFE ^52^. Another gene with shorter distance to known epilepsy genes, *FLNA*, has itself been associated with epilepsy and seizure disorders ^5^. *FLNA* is also known to interact with *HCN1* channels during neuronal excitability modulation in the mature brain ^53^. Additionally, *FLNA* also controls ECM remodeling by regulating metalloproteinase activity and hence ECM degradation ^54^. Based on the enrichment of ECM genes, we suggest that especially for NAFE, genetic variants which may impact ECM morphology could lead to imbalance in excitatory and inhibitory signals ^55^ and hence underlie epileptogenesis.

Consistent with the assumed pathomechanisms, we identify a significant enrichment of QRVs in genes related to brain development only in DEE (Fig. 2) and GGE (Fig. 1). Interestingly, most genes with high developmental scores are assigned to DEE (Fig. 3), suggesting that rare variants in these genes may contribute to the disease development. *TNC*, which encodes an ECM protein, was identified in both DEE and GGE (Fig. 1). It controls neurite growth and axon guidance ^56^ and it is highly active during early brain development, which is mirrored by a high developmental score (Fig. 3). Intriguingly, *TNC* is higher expressed by both neurons and glias after seizures, which can lead to ECM remodeling and induce additional epileptic events ^56,57^. For NAFE, genes with high developmental score cluster in the inferolateral temporal cortex (Fig. 2), a signal that may be triggered by a high number of temporal lobe epilepsy cases within the NAFE cohort.

Aside from the established list of epilepsy associated genes from the Epi25 cohort, there are a number of curated lists for epilepsy genes, for instance the SAGAS database, containing candidate genes with possible polygenic and monogenic causal tendencies ^5^ and Genes4Epilepsy (https://github.com/bahlolab/Genes4Epilepsy). We identified a significant overlap of the QRVs genes from our study with both SAGAS (DEE: pval = 0.001, NAFE: pval = 1.30e-8, GGE: pval = 0.02) and Genes4Epilepsy (NAFE: pval = 0.001), which lends additional support to our findings.

A previous study of the Epi25 Collaborative suggested that clinical presentations of GGE and NAFE are influenced by common and rare variants, as opposed to DEE which is mainly caused by *de novo* ultra-rare highly deleterious variants ^11^. Here, we focused on rare variants, present in controls, but at higher OR in epilepsy patients. Our results support the hypothesis that rare variants could be important in NAFE pathomechanism. Moreover, ECM appears to play a central role in NAFE. For DEE we retrieve genes that have high expression during development, which meets the expected pathomechanism; however, the number of identified genes is rather low. Based on the genes identified for GGE we cannot infer which pathways play an important role in the pathomechanism. It is possible that either enlarged patient cohorts or a focus on common variants will shed more light on GGE ^58^ pathophysiolog.

## Supporting information

GO Results

Variant Enrichment Results

Custom ORA

## Data Availability

The data that supports this study are available in https://epi25.broadinstitute.org/downloads. The code used in the analysis is deposited under https://github.com/lbundalian/EPI25_VSEA.

https://github.com/lbundalian/EPI25_VSEA

## 8. Acknowledgement

Open Access funding was enabled and organized by Projekt DEAL. This study is funded by the Else Kröner-Fresenius-Stiftung 2020_EKEA.42 to D.L.D. and the German Research Foundation SFB 1052 project B10 to D.L.D. and A.G., D.L.D. is funded through the “Clinician Scientist Programm, Medizinische Fakultät der Universität Leipzig”. We thank the Epi25 principal investigators, local staff from individual cohorts, and all of the patients with epilepsy who participated in the study for making possible this global collaboration and resource to advance epilepsy genetics research. This work is part of the Centers for Common Disease Genomics (CCDG) program, funded by the National Human Genome Research Institute (NHGRI) and the National Heart, Lung, and Blood Institute (NHLBI). CCDG-funded Epi25 research activities at the Broad Institute, including genomic data generation in the Broad Genomics Platform, are supported by NHGRI grant UM1 HG008895 (PIs: Eric Lander, Stacey Gabriel, Mark Daly, Sekar Kathiresan). The Genome Sequencing Program efforts were also supported by NHGRI grant 5U01HG009088-02. The content is solely the responsibility of the authors and does not necessarily represent the official views of the National Institutes of Health. We thank the Stanley Center for Psychiatric Research at the Broad Institute for supporting the genomic data generation efforts.

## 9. Author Contribution

**L.B**. Conceptualization; writing – original draft; formal analysis; investigation; methodology. **D.L.D**. Investigation; methodology. **S.C**., **A.V**., **F.B**., **D.L**., **H.O.H** Methodology; validation; writing – original draft. **Y-Y.S**. and **C–C.L**. Methodology; formal analysis; writing – original draft. **A-S. K** and **A.G** Conceptualization; writing – original draft. **J.R.L**. and **D.L.D**. Conceptualization; writing – original draft; supervision; funding acquisition.

## 10. Ethical declaration

This study was approved by the ethics committee of the University of Leipzig, Germany (224/16-ek and 402/16-ek) and by the Epi25 Strategy Committee (approval from 16.10.2019). The availability of informed consent from the tested individuals was checked as part of Epi25 sample inclusion criteria. Since data analysis was performed across multiple centers, we used only aggregated data to assure patient anonymity.

## Supplemental Figures

**Figure S1.**
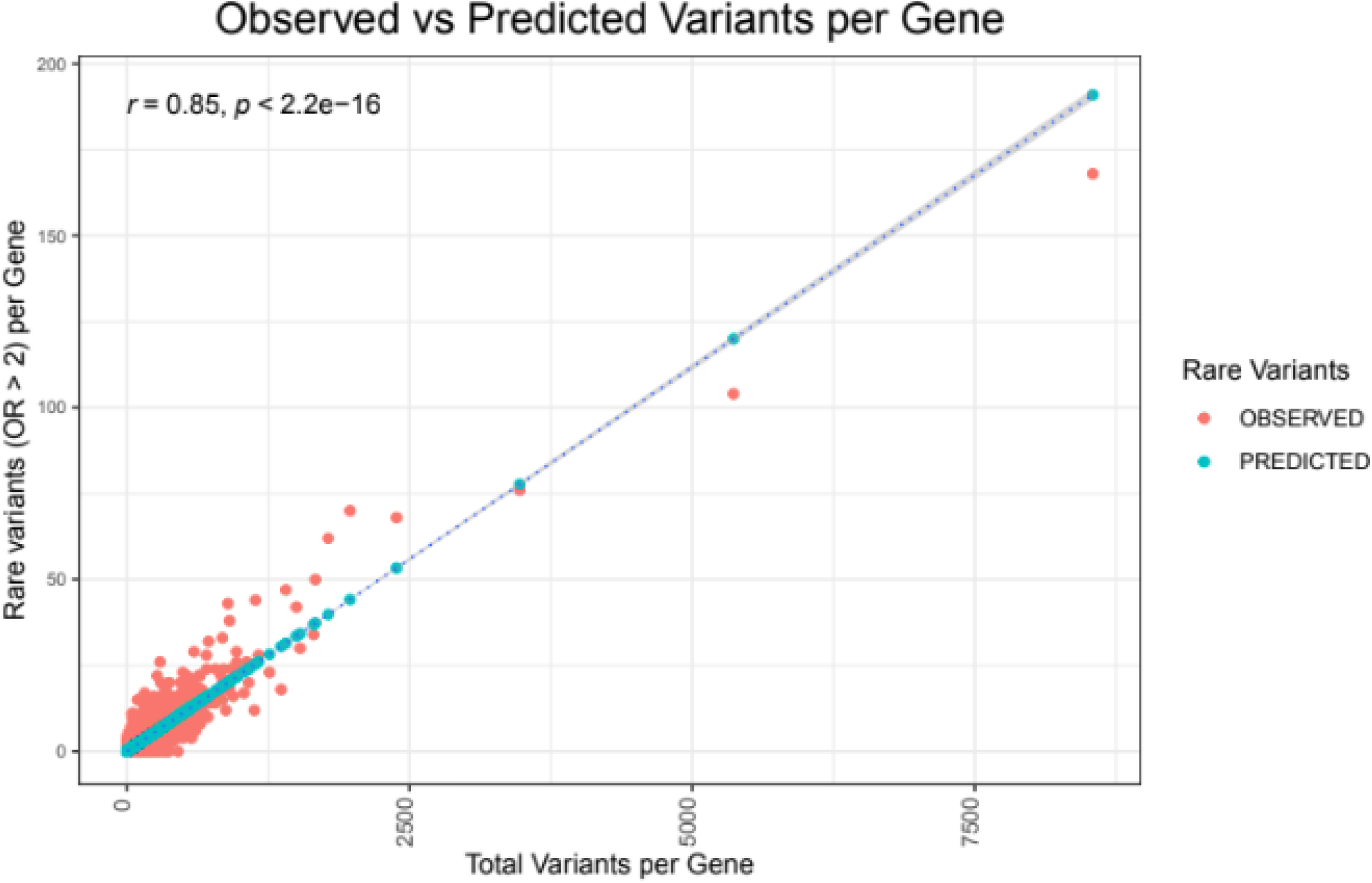
Relationship of the number of rare variants and total number of variants in a gene. The linear regression model (r = 0.85) established that for a given number of variants per gene (total variants), we can estimate the expected number of rare variants. Only those genes which had an excess of expected number variants (i.e. above the dotted slope) were considered for the VSEA.

**Figure S2.**
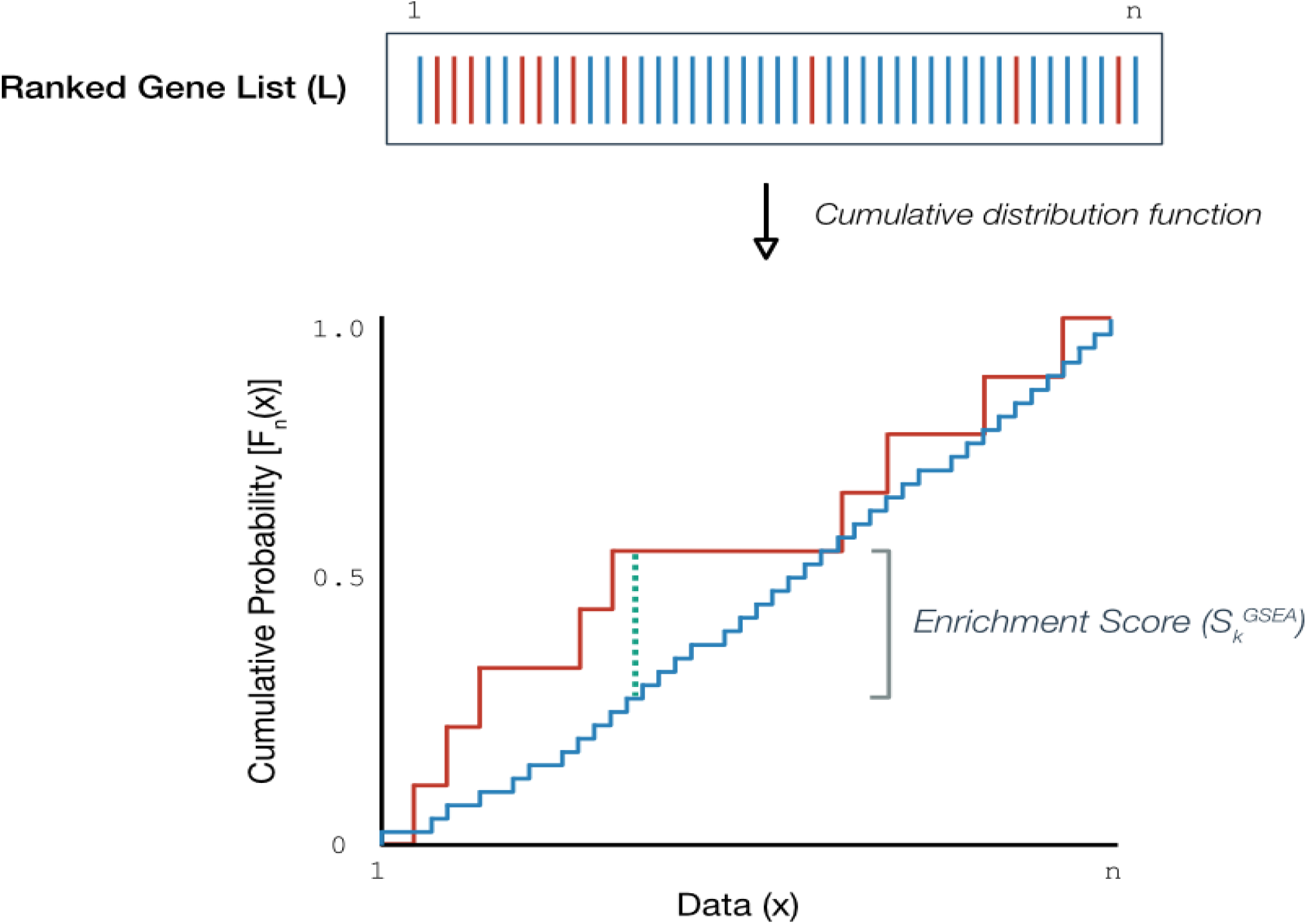
Cumulative ranked sum for hits and miss of QRVs. For every QRVs found in the list of variants having a higher frequency in patients – Data(x), the cumulative HIT score (red) will increment by one otherwise the cumulative MISS score (blue) will be incremented. The normalized difference between the MISS and HIT score yields the ENRICHMENT SCORE. (https://www.pathwaycommons.org/guide/primers/data_analysis/gsea/#safe)

**Figure S4.**
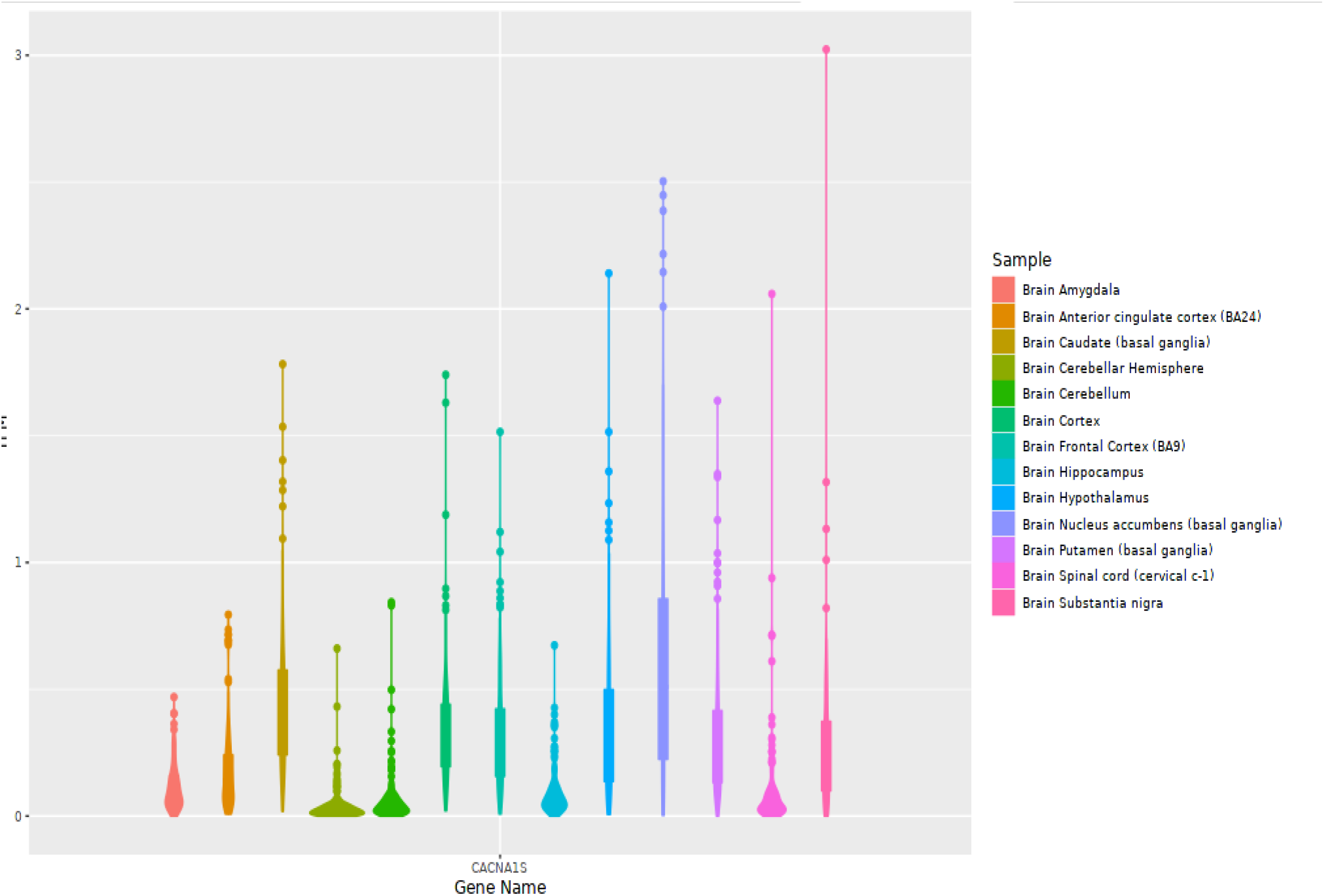
Expression of *CACNA1S* from Single Gene Analysis. *CACNA1S* is not that highly expressed in brain. However, using the *Single Gene Analysis* feature of PTEE (https://bioinf.eva.mpg.de/PTEE), we can observe the outliers in across different brain-related tissues. A previous study suggested that the voltage-gated calcium channel encoded by *CACNA1S* could play a role in mediating calcium influx and neuronal excitability (*Cain, S. M*., *& Snutch, T. P. (2012). Voltage-Gated Calcium Channels in Epilepsy. In J. L. Noebels (Eds*.*) et. al*., *Jasper’s Basic Mechanisms of the Epilepsies. (4th ed*.*). National Center for Biotechnology Information (US)*)

